# Ultralow-field portable MRI feasibility and safety in pediatric and neonatal ECMO: a single center year-long experience

**DOI:** 10.1101/2025.05.12.25327194

**Authors:** Jessica S Wallisch, Asdis Finnsdottir Wagner, John M Daniel, Allison Taber, Maura Sien, Sarah Foster, Nathan Artz, Jose A Pineda, Patrick M Kochanek, Sherwin S Chan

**Affiliations:** Division of Pediatric Critical Care Medicine, Department of Pediatrics, Children’s Mercy Hospital, Kansas City, MO; University of Missouri-Kansas City School of Medicine, Kansas City, MO; Division of Neonatology, Department of Pediatrics, Children’s Mercy Hospital, Kansas City, MO; Department of Radiology, Children’s Mercy Hospital, Kansas City, MO; Division of Critical Care, Department of Pediatrics, DGSOM at UCLA; Department of Critical Care Medicine, Safar Center for Resuscitation Research, School of Medicine, University of Pittsburgh, Pittsburgh, PA

**Author notes:** <u>Corresponding author</u>: Jessica S. Wallisch, MD, Children’s Mercy Hospital, 2401 Gillham Road, Kansas City, MO 64108.

## Abstract

**Introduction:** Extracorporeal membrane oxygenation (ECMO) outcomes continue to improve, yet high rates of acute brain injury (ABI) threaten survival with significant morbidity for ECMO survivors. Currently available imaging modalities [ultrasound and computed tomography (CT)] have low early detection rates for hypoxic-ischemic and cerebrovascular injuries, delaying the diagnosis of ABI while on ECMO. CT sensitivity increases only when it may be too late to effectively intervene. High-field (>1.5 Tesla) magnetic resonance imaging (MRI), the gold standard to diagnose ischemic brain injury, is not compatible with ECMO. An FDA-cleared ultralow-field (0.064 Tesla) portable MRI (pMRI) has been studied in diverse types of ABI and with equipment that is typically not MRI compatible. There is very limited experience using pMRI in ECMO and even less in pediatric ECMO patients, therefore additional feasibility and safety data is needed in this cohort.

**Methods:** This single center, IRB approved study was conducted at a free-standing quaternary children’s hospital. All neonatal and pediatric patients cannulated onto ECMO were screened for eligibility. Subjects underwent bedside ultralow-field pMRI with Swoop (Hyperfine, Guilford, CT). Data on ECMO variables, time for patient positioning and scan, MRI sequences, concurrent critical care equipment, changes in ECMO flow and vital signs, and cannula displacement was collected.

**Results:** Over a 1-year period (Aug 2023-Aug 2024) 41 patients were screened. 16 out of 20 enrolled subjects had pMRI attempted and 13 (81.25%) received the full imaging protocol (T1, T2, FLAIR and DWI). The median staff members for pMRI positioning was 6 [5, 7] compared to 7 [7,7] for head CT (0.03). The median positioning and pMRI imaging time was 66 min [56, 70] compared to 75 min [70,79] for intrahospital transport for head CT. One subject had a ≥ 20% decrease in mean arterial pressure, however remained within the clinical goals without intervention. Unlike during head CT imaging acquisition, continuous renal replacement therapy was not interrupted during pMRI.

**Conclusions:** pMRI is safe and feasible in pediatric ECMO with no clinically relevant complications seen in our cohort. Resource utilization and delivery of concurrent critical care is superior for bedside imaging compared to intrahospital transport to CT.

**Clinical Trial Registration:** NCT06074406 https://clinicaltrials.gov/study/NCT06074406?term=NCT06074406Crank=1

**CLINICAL PERSPECTIVE:** *What is new?:*

- This study expands the experience of ultralow-field portable MRI to pediatric ECMO patients.
- Ultralow-field portable MRI is feasible and less resource-intense to perform on pediatric ECMO patients compared to intrahospital transport for head CT.
- Pediatric ECMO patients tolerate bedside MRI without clinically significant changes in ECMO flows, perfusion, or oxygenation.

**What are the clinical implications?:**

- Ultralow-field portable MRI can expand time-sensitive head imaging options for pediatric patients on ECMO with less interruptions of critical care therapies, decreased resource utilization, and eliminated risks of travel and radiation exposure.
- Timely diagnosis of acute brain injury while on ECMO can prompt changes in neuromonitoring, anticoagulation management, and delivery of neuroprotective care with the intent to improve neurologic outcomes of pediatric ECMO survivors.

## INTRODUCTION

Significant advances in extracorporeal membrane oxygenation (ECMO) management and technology have expanded its use and improved survival rates, yet rates of poor neurologic outcomes remain unchanged following this level of support^1–7^. There are considerable risks to the development of acute brain injury (ABI) in this population across the continuum of care (before ECMO, during cannulation, and during ECMO support) due to a myriad of patient, disease, and treatment factors. Hypoxia, hypotension, and cardiac arrest (which occurs in 40% of pediatric ECMO patients) are among the known factors implicated in the pre-ECMO period with alterations in cerebral blood flow, thromboembolic events, and use of systemic anticoagulation being well described risks during ECMO support. The totality of these risk factors results in half of pediatric ECMO survivors having poor neurologic outcomes and disability^5,8^.

There are different types of ABI that occur in this population including hypoxic-ischemic, cerebrovascular (stroke), and hemorrhagic injuries. The ability of clinicians to diagnose these different types of ABI is limited by multiple factors including 1) most pediatric ECMO patients require sedation — limiting the neurologic examination, 2) significant limitations to noninvasive neuromonitoring, 3) risks of invasive neuromonitoring, and 4) lack of sensitivity in available imaging modalities for early hypoxic-ischemic and cerebrovascular type injuries. Head ultrasound is one imaging modality that can be easily obtained at the bedside in patients younger than 1 year old with an open fontanelle, but ultrasound tends to miss ischemic and posterior fossa injuries with only a 41% sensitivity (94% positive predictive value and 36% negative predictive value) for ABI on ECMO^9^. CT can be used across all ages of pediatric patients on ECMO but adds significant risks and logistical challenges related to intrahospital transport and uses ionizing radiation^10–12^. Furthermore, CT’s detection of early ischemia is inferior, with only 27.3% sensitivity in acute pediatric arterial ischemic stroke, which leads to delays in management^13,14^. While some institutions have access to portable CT, this does not increase its diagnostic accuracy for all types of ABI on ECMO nor eliminate risks related to radiation exposure. Traditional MRI is the gold-standard to diagnose hypoxic-ischemic and cerebrovascular injuries, but it is incompatible with ECMO^15^. It is estimated that less than half of pediatric ECMO patients receive head imaging with CT or a post-decannulation MRI, yet 21-60% of those who receive imaging have acute neuroimaging abnormalities^8,16,17^. This leaves clinicians with diagnostic uncertainty during a critical time window, when ABI is suspected, resulting in delayed diagnosis and missed opportunity for neuroprotective or directed interventions^18–20^. Patients might also receive unnecessary empiric treatment that also carries risks such as hyperosmolar therapy or changes to anticoagulation management in cases of suspected ABI without neuroimaging confirmation.

Ultralow-field portable MRI (pMRI) is a newer FDA cleared imaging modality that has fewer patient support equipment limitations, eliminates ionizing radiation exposure and the need for transport, and experience with its use is rapidly expanding in adult ABI including after cardiac arrest, stroke, and intracerebral hemorrhage^21–25^. However, there is limited experience in pediatric patients because of the need for programming modifications when imaging the developing brain (i.e. differences in brain-tissue composition in neonates)^26,27^. There is even more limited published experience with the use of ultralow-field pMRI in ECMO with only two small adult case series and a multicenter prospective observational cohort study of 50 adult patients^28–30^. Pediatric experience lags with one reported case series of 4 patients^31^. The primary aim of this study was to expand the feasibility and safety data for use of pMRI in pediatric ECMO patients.

## METHODS

### Study Design

This was a single center cohort study at a quaternary free-standing children’s hospital with high ECMO volume and ELSO Platinum Center of Excellence designation. Neonates and children (age 0 to <18 years) admitted to the Neonatal Intensive Care Unit (NICU), Pediatric Intensive Care Unit (PICU), or Cardiac Intensive Care Unit (CICU) and receiving venovenous (VV) or venoarterial (VA) ECMO support between September 2023 and August 2024 were prospectively screened and enrolled. Patients were excluded for pregnancy, active implants (permanent pacemaker or defibrillator, deep brain stimulator, cochlear implant, vagal nerve stimulator, programmable shunt), and MRI incompatible surgical hardware or shrapnel. The study was approved by the Institutional Review Board of Children’s Mercy Hospital (STUDY00002824).

### Study Procedure

Ultralow-field portable MRI imaging was completed with Swoop (device version 1.8 with software 8.7 beta, Hyperfine, Guilford, CT) which has a static magnetic field strength of 0.064 Tesla (T). The imaging protocol included T1-weighted, T2-weighted, Fluid Attenuated Inversion Recovery (FLAIR), and Diffusion Weighted Imaging (DWI with calculated Apparent Diffusion Coefficient) using the standard product sequences excepting modified sequences for patients < 1 year. Scanning procedures were coordinated with bedside staff and ECMO teams to avoid clinical care interference with goal scan time occurring within 72 hours of ECMO cannulation as this would capture injuries occurring in the pre-cannulation, peri-cannulation, and immediate post-cannulation periods where the largest changes occur in cerebral blood flow and oxygenation. Radiology research coordinators who are trained radiology technologists transported the pMRI to the bedside of enrolled subjects. Subjects were positioned into the pMRI head coil by a team of bedside staff (critical care nurses, respiratory therapists, intensivists) and ECMO team members (perfusionists and specialists). The ECMO team monitored vital signs, flows, oxygenation, and support devices per clinical protocol for the duration of the scanning procedure.

### Data Collection

Data was collected on patient variables (age and weight at cannulation, sex, race and ethnicity, ICU location, survival), ECMO variables (type and indication for ECMO, cannula type and location, pump type and circuit size, cannulation and decannulation dates), and pMRI variables (rate of scan completion, reason for lack of completion, timing of scan, sequences run, reason exam ended early if applicable). Data was also collected on imaging procedure metrics (positioning/prep time, scan duration, total imaging procedure time, number of staff required for positioning, concurrent ICU therapies/devices), rates of adverse or safety events, and if head CT was obtained by the medical team during the study period. Data was recorded and stored in REDCap (v14.6.8).

### Outcomes

The primary feasibility outcome was defined as rate of imaging completion with all needed sequences (T1, T2, FLAIR and diffusion weighted imaging) relative to scans attempted. Secondary feasibility outcomes included total time for positioning and imaging, the number of staff required for positioning, and rates of concurrent therapies that were paused. Primary safety outcomes were modeled after the SAFE-MRI ECMO study^29^ and defined as change in ECMO cannula positioning requiring repositioning by surgeon, ≥20% decrease in ECMO flow, ≥ 20% decrease in mean arterial pressure (MAP), or ≥ 10% in pulse oximetry (SpO_2_).

### Statistical Analysis

Descriptive statistics were reported as median with interquartile range and frequency (%). Analysis was conducted in SPSS (IBM, v29).

## RESULTS

### Patient Demographics and ECMO Details

All patients cannulated onto ECMO at Children’s Mercy Hospital over a one-year period beginning August 2023 (n=41) were screened for eligibility. There were 19 unique subjects consented and enrolled that comprised 20 total ECMO courses (Figure 1). The median age of our cohort was 4.21 months [0.16, 5.85] with a median weight of 4.55 kg [3.20, 7.91]. Half of the subjects enrolled were admitted to the CICU. The patient demographics are summarized in Table 1.

**Figure 1.**
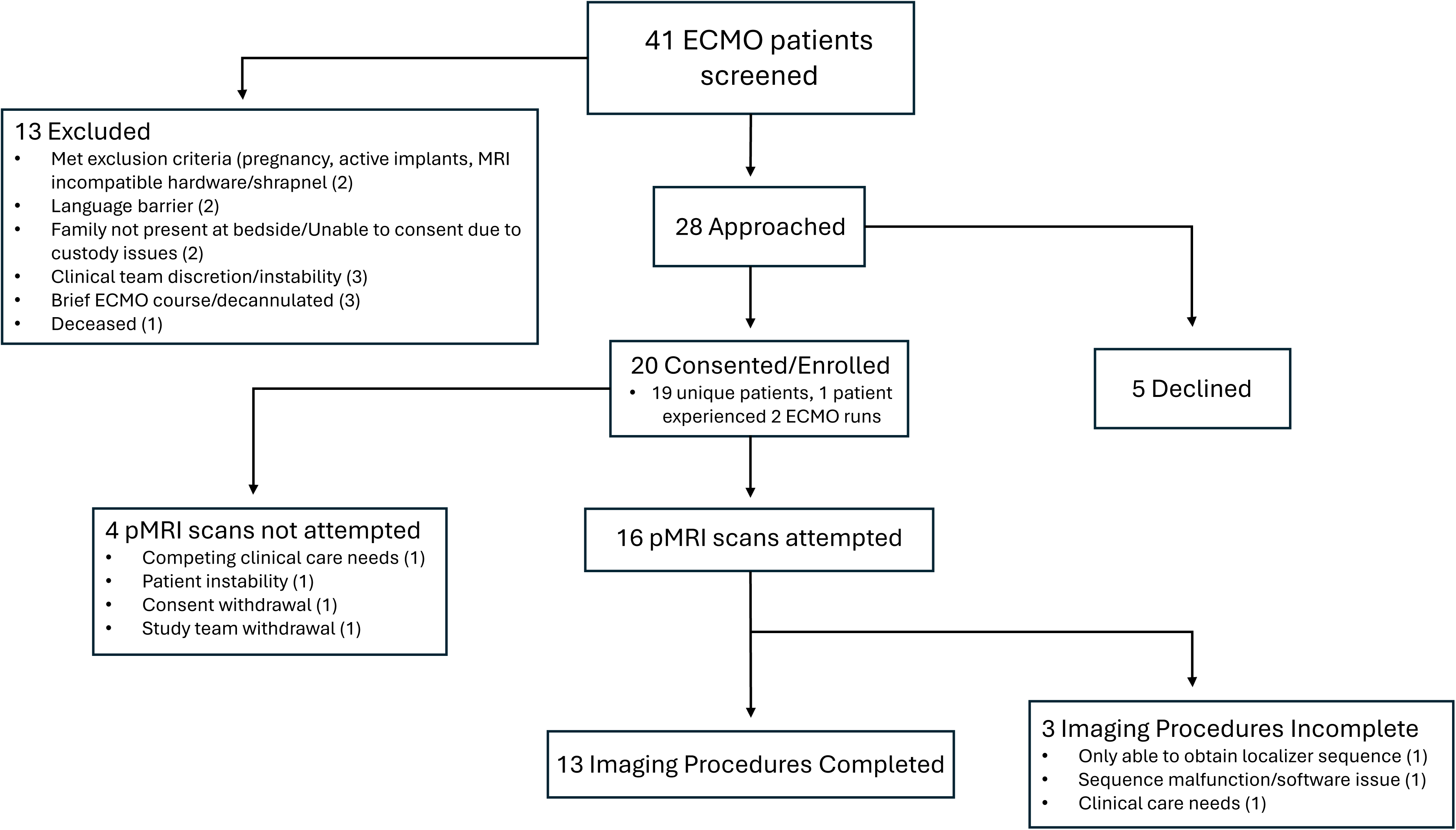
Consort flow diagram of patient selection. ECMO, extracorporeal membrane oxygenation; pMRI, portable magnetic resonance imaging.

**Table 1.** Patient Demographics, ECMO variables and Clinical outcomes. Abbreviations: Pediatric Intensive Care Unit (PICU), Cardiac Intensive Care Unit (CICU), Intensive Care Nursery (ICN), extracorporeal membrane oxygenation (ECMO), venovenous (VV), venoarterial (VA), extracorporeal cardiopulmonary resuscitation (eCPR)

|  | ECMO Patients |  |
| --- | --- | --- |
|  | N (%) | Median [IQR] |
| **Location** |  |  |
| PICU | 4 (20%) |  |
| CICU | 10 (50%) |  |
| ICN | 6 (30%) |  |
| **Demographics** |  |  |
| Age (months) |  | 4.21 [0.16, 5.85] |
| Weight (kg) |  | 4.55 [3.20, 7.91] |
| Male | 17 (85%) |  |
| **Race** |  |  |
| Caucasian | 12 (60%) |  |
| Black or African | 2 (10%) |  |
| More than one | 4 (20%) |  |
| Unknown | 2 (10%) |  |
| **Ethnicity** |  |  |
| Non-Hispanic | 16 (80%) |  |
| Hispanic | 1 (5%) |  |
| Unknown | 3 (15%) |  |
| **ECMO type** |  |  |
| VV | 3 (15%) |  |
| VA | 17 (85%) |  |
| **ECMO indication** |  |  |
| Respiratory | 9 (45%) |  |
| Cardiac | 4 (20%) |  |
| eCPR | 7 (35%) |  |
| **ECMO pump type** |  |  |
| Roller | 16 (80%) |  |
| Centrifugal | 4 (20%) |  |
| **Cannulation site** |  |  |
| Neck | 16 (80%) |  |
| Chest | 4 (20%) |  |
| **Early clinical outcomes** |  |  |
| 30-day ECMO Survival | 15 (79%) |  |
| ICU Survival | 14 (74%) |  |

A majority (85%) of the cohort was cannulated onto VA ECMO with 80% having neck cannulation sites. The indications and other ECMO details are summarized in Table 1. The median ECMO duration of our cohort was 177 hours [127, 371]. A total of 15 subjects (78.95%) survived greater than 30 days following ECMO decannulation with 14 subjects (73.68%) surviving to ICU discharge.

### pMRI Feasibility

Imaging with pMRI was attempted in 16 out of 20 enrolled subjects (80%). Reasons imaging was not attempted included competing clinical needs (1), patient instability (1), study withdrawal prior to imaging (1), and study team withdrawal due to need for safety clarification for temporary pacing wires (1). The majority of subjects in which a scan was attempted (9, 56.25%) were scanned less than 72 hours following cannulation with 5 subjects (31.25%) completing pMRI less than 24 hours from cannulation. The median ECMO duration at the time of pMRI was 41.83 hours [19.78, 106.43]. Thirteen out of 16 (81.25%) subjects were able to complete the full imaging sequence protocol. A sample of acquired images is provided in Figure 2. Two subjects only had a localizer image obtained and reasons for early imaging termination included a sequence malfunction during diffusion weighted imaging acquisition (1) and patient return to the cardiac catheterization laboratory for reasons unrelated to pMRI (1).

**Figure 2.**
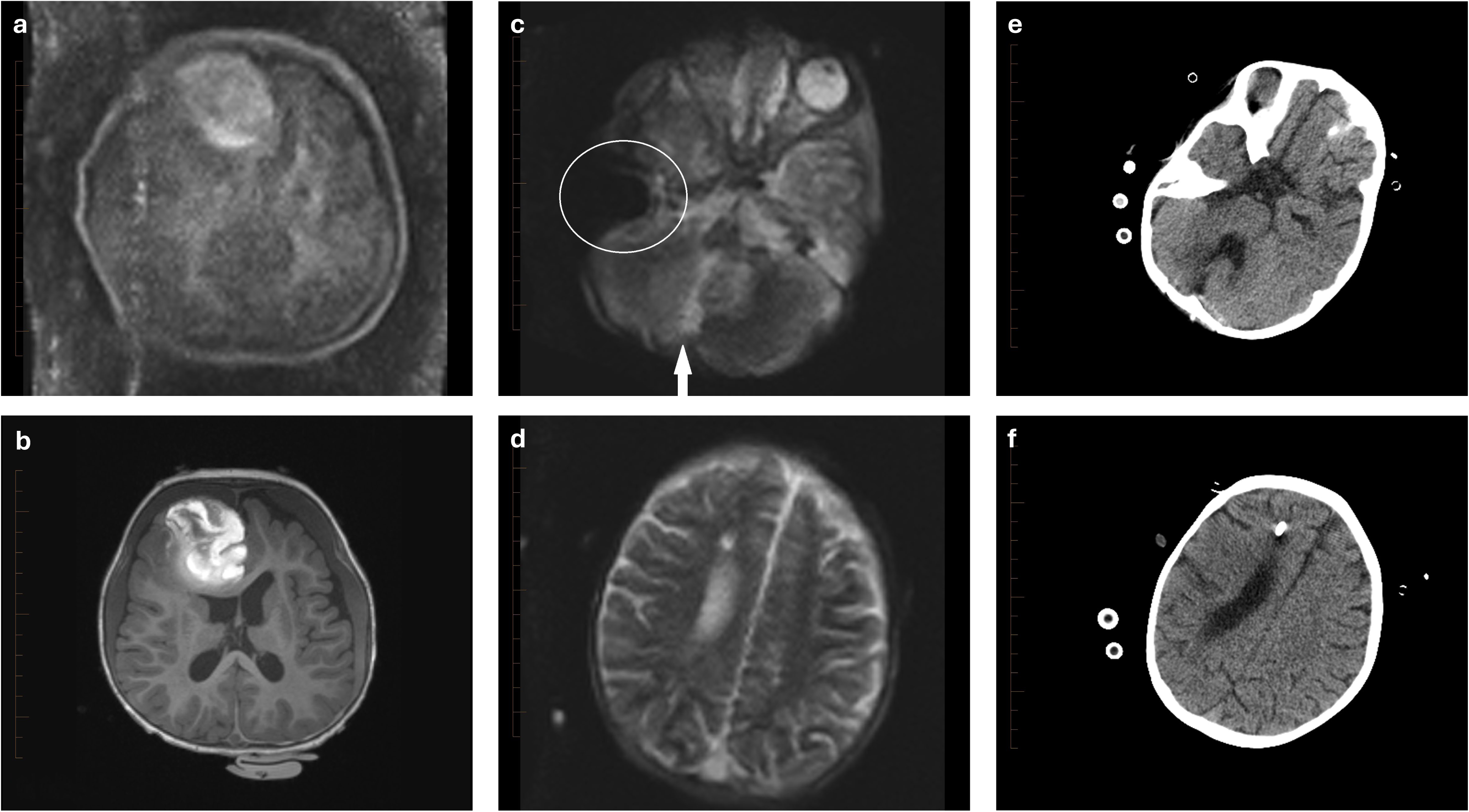
Representative images acquired by pMRI with comparison head imaging on pediatric ECMO patients. (a-b) Brain imaging of a toddler age subject with intraparenchymal hemorrhage in the right frontal lobe. (a) Axial T1 image from the ultralow-field pMRI. (b) Axial T1 image from a standard of care high-field MRI performed 6 days prior to the pMRI. (c-f) Brain imaging of a toddler age subject who underwent resection of a posterior fossa tumor and placement of a right frontal approach intraventricular catheter. (c) Axial T2 image from the ultralow-field pMRI showing some of the post-surgical changes around the resection cavity (arrow). Also note the signal dropout from the ECMO catheter (circle). (d) A more superior axial T2 image shows the course of the intraventricular catheter. Axial CT images performed 5 days after the pMRI show the resection changes (e) and the catheter tract (f). pMRI, portable magnetic resonance imaging; ECMO, extracorporal membrane oxygenation; CT, computed tomography.

The median time for positioning was 23 min [16, 24] with median pMRI scan duration of 33 min [31.50, 39.50]. The median number of staff present was 6 [5, 7] with 5 [4, 5] actively involved in positioning the patient for pMRI. The only concurrent critical care monitoring or therapy paused during pMRI was continuous electroencephalogram (EEG) monitoring due to interference; however, MRI-compatible leads remained on the patient minimizing the duration off monitoring (Table 2).

**Table 2.**
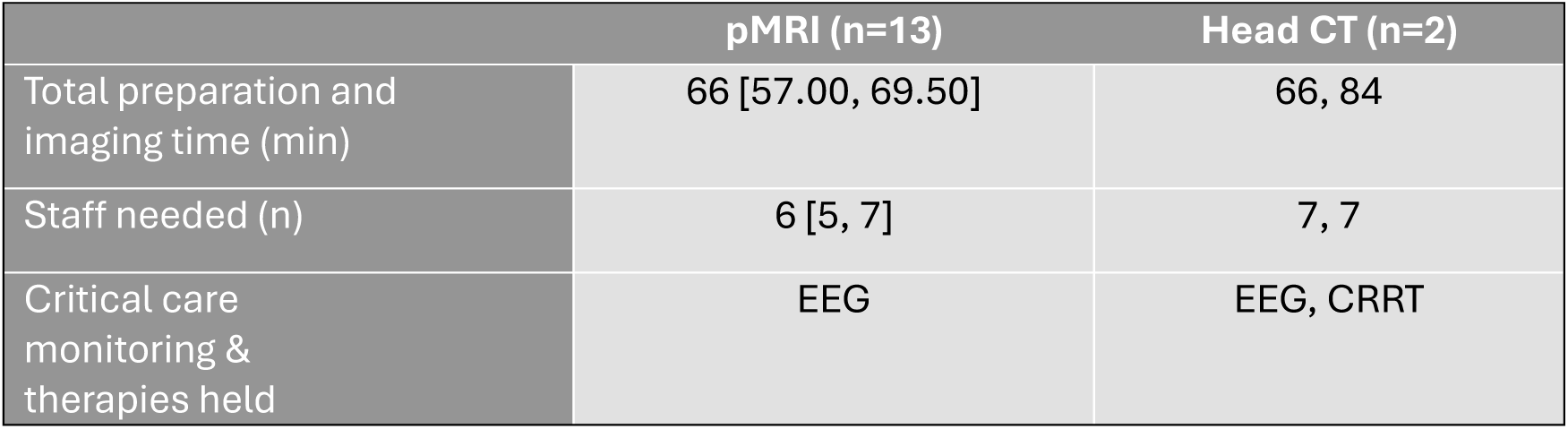
Portable MRI Outcomes Relative to CT. Values are expressed as median [IQR]. Abbreviations: minutes (min), portable MRI (pMRI), computed tomography (CT), electroencephalogram (EEG), continuous renal replacement therapy (CRRT)

### Safety

During image acquisition one patient had a predefined safety event with ≥20% decline in MAP, but still remained within the clinical goals and did not require intervention or early study procedure termination. One patient required ECMO arterial cannula repositioning 8 days following pMRI, however the cannula positioning was stable on subsequent chest x-ray and echocardiography obtained later on the same day after pMRI imaging. No other adverse events or serious adverse events were attributed to study interventions.

### pMRI relative to CT

Half of the patients (10, 50%) traveled to CT during study enrollment. A majority (7, 70%) of CT scans were obtained for head imaging followed by chest (2, 20%) then abdomen/pelvis (1,10%). Two of the head CTs (28.57%) were completed during study coordinator working hours and available for tracking. The median time outside the ICU was 26.5 minutes [21, 32] with median total preparation, imaging, and travel time of 75 minutes [66, 84] (Table 2). Both head CTs utilized 7 staff members, which was more resource-intense compared to pMRI. Two subjects had continuous renal replacement therapy (CRRT) interrupted and one patient had their EEG leads temporarily removed prior to travel for CT.

## DISCUSSION

This study completed pMRI in the largest cohort of pediatric ECMO patients to date. We demonstrated expanded feasibility with similar preparation plus imaging time and less resource-intensity including staff members with lack of transport out of the intensive care unit as compared to the currently clinically available modality of head CT. Additionally, a majority (81.2%) of imaging attempts were completed with all planned sequences. Imaging was deemed safe with no subjects having clinically relevant changes to ECMO flows, MAP, or oxygenation during study procedures nor did we experience any migration of ECMO cannulas attributable to study interventions.

Our study evaluated similar safety metrics to the SAFE MRI ECMO study which evaluated the use of pMRI in 50 adult ECMO patients^28,32^. With the exception of a non-clinically relevant decline in MAP, subjects in our smaller cohort did not experience any adverse events or pre-defined safety outcomes attributable to study interventions unlike the 3 (6%) reported by Cho et al. One subject required an arterial cannula repositioning which was deemed unrelated to study interventions as it occurred 8 days after pMRI imaging with evidence of stable cannula positioning on x-ray and echocardiography immediately following the study intervention. It was noted that this subject underwent intrahospital transport to obtain a head CT on the day prior to observing a change in arterial cannula position via x-ray which was then confirmed on echocardiography. While there is a temporal association with intrahospital transport, the true etiology or etiologies of the cannula displacement could not be determined. Nevertheless, as this is a known complication of intrahospital transport on ECMO and this requires a much larger patient and circuit movement, it could be hypothesized that the risks of intrahospital transport are greater than those for pMRI.

Adult and pediatric ECMO populations have different risk factors and patterns of ABI, cannulation sites, and brain water content requiring modifications in the approach to imaging. The only prior published experience with pMRI in pediatric ECMO was in 4 peripherally cannulated patients with no adverse events while obtaining only T1 and T2 weighted sequences, resulting a shorter scan duration without the ability to detect acute ischemia^31^. Therefore, while the work of Sabir et al was fundamental to exploring the use of pMRI in pediatric ECMO, our study built upon their findings by increasing the cohort size, using a more complete pMRI protocol that included DWI, additional granularity of feasibility and safety outcomes, and comparison to CT imaging.

We met our goal to obtain imaging with pMRI within 72 hours of cannulation in a majority of subjects (56.25%) with a median time to scan of 41.83 hours. We identified this goal timeframe due to the vulnerability and higher risk of acute brain injury during the pre- and peri-cannulation time periods^2,33–36^. As we prioritized clinical care needs during this feasibility pilot, the scan time was subject to downtime between patient care, coordination and availability of all team members required to move a patient on ECMO, and the research coordinators’ work hours and other responsibilities. Nonetheless, the fact that 31.25% of imaged subjects’ scans were obtained in under 24 hours from cannulation provides proof of concept that pMRI could be utilized to better delineate and ascribe patterns of ABI within the timeframe of high risk for ABI in this patient population. Our feasibility data will inform future efforts to determine optimal timing of pMRI imaging and guide implementation strategies.

As this was a single center cohort study conducted over 1 year, our cohort size is limited. Our institution averages 40-50 ECMO runs annually, thus the number of subjects screened was typical. Our enrollment was limited by not only exclusion criteria but also inability to approach families/surrogate decision makers due to language barriers (2), timing of presence at the bedside (2), clinical instability (2), and rapid decannulation or death (3). Our study team’s effort to avoid interrupting clinical care and the standard research coordinator work hours impacted the timing of scan attempts and completion. Many of these limitations would not be a factor if this imaging modality becomes standard of care.

Our study cohort had a high short-term survival rate with overall all-cause mortality of 26.32%, which is consistent with our center ELSO registry data of 29.70%, however could skew our findings toward less severe ABI. Notably our institution had 3 ECMO patients declared deceased by neurologic criteria (1.7%), a total of 23 patients with ischemic brain injury (12.7%), and a total of 21 patients with hemorrhagic brain injury (11.6%) over the same 3-year span as the reference mortality data (2020 – 2023). As our feasibility study did not assess imaging findings or rate of acute brain injury detection, this limitation did not impact the interpretation of our results. Finally, this pilot study was not powered for safety outcomes. At a 7.7% cannula displacement prevalence in our cohort, we calculate the 95% CI to be [1.2%, 36.0%]

## CONCLUSION

pMRI is feasible and less resource-intense than intrahospital transport to CT for pediatric ECMO patients. Future work will expand safety data and determine the diagnostic yield of ultralow-field portable MRI for ischemic and hemorrhagic acute brain injuries in pediatric ECMO patients. This has the potential to significantly improve neuroimaging and neurocritical care for pediatric ECMO patients by allowing for earlier and more accurate diagnosis, better-informed decision making and treatment, and ultimately better neurologic outcomes and survival.

## Data Availability

Data is available upon reasonable request.

## Non-standard Abbreviations and Acronyms

ECMO: extracorporeal membrane oxygenation
ABI: acute brain injury
CT: computed tomography
MRI: magnetic resonance imaging
pMRI: portable magnetic resonance imaging
FDA: Food and Drug Administration
T: Tesla
VV: venovenous
VA: venoarterial
ELSO: Extracorporeal Life Support Organization
ICN: Intensive Care Nursery
PICU: Pediatric Intensive Care Unit
CICU: or Cardiac Intensive Care Unit
FLAIR: Fluid-attenuated inversion recovery
DWI: diffusion-weighted imaging
EEG: electroencephalogram
CRRT: continuous renal replacement therapy

## ACKNOWLEDGMENTS

The authors would like to thank Hung-Wen Yeh PhD, Division of Health Services and Outcomes Research, Children’s Mercy Research Institute, Kansas City, MO, for statistical consultation services.

## REFERENCES

1. Alsoufi B, Al-Radi OO, Nazer RI, Gruenwald C, Foreman C, Williams WG, Coles JG, Caldarone CA, Bohn DG, Van Arsdell GS. Survival outcomes after rescue extracorporeal cardiopulmonary resuscitation in pediatric patients with refractory cardiac arrest. The Journal of thoracic and cardiovascular surgery. 2007;134:952–959.e952. doi: 10.1016/j.jtcvs.2007.05.054

2. Barrett CS, Bratton SL, Salvin JW, Laussen PC, Rycus PT, Thiagarajan RR. Neurological injury after extracorporeal membrane oxygenation use to aid pediatric cardiopulmonary resuscitation. Pediatric critical care medicine : a journal of the Society of Critical Care Medicine and the World Federation of Pediatric Intensive and Critical Care Societies. 2009;10:445–451. doi: 10.1097/PCC.0b013e318198bd85

3. Cho SM, Canner J, Chiarini G, Calligy K, Caturegli G, Rycus P, Barbaro RP, Tonna J, Lorusso R, Kilic A, et al. Modifiable Risk Factors and Mortality From Ischemic and Hemorrhagic Strokes in Patients Receiving Venoarterial Extracorporeal Membrane Oxygenation: Results From the Extracorporeal Life Support Organization Registry. Critical care medicine. 2020;48:e897–e905. doi: 10.1097/ccm.0000000000004498

4. Cook RJ, Rau SM, Lester-Pelham SG, Vesper T, Peterson Y, Adamowski T, Sturza J, Silverstein FS, Shellhaas RA. Electrographic Seizures and Brain Injury in Children Requiring Extracorporeal Membrane Oxygenation. Pediatric neurology. 2020;108:77–85. doi: 10.1016/j.pediatrneurol.2020.03.001

5. Dhar AV, Scott S, Anton-Martin P, Tweed J, Morris MA, Modem V, Raman L, Golla S. Neurodevelopmental Outcomes in Extracorporeal Membrane Oxygenation Patients: A Pilot Study. ASAIO journal (American Society for Artificial Internal Organs : 1SS2). 2020;66:447–453. doi: 10.1097/mat.0000000000001035

6. Madderom MJ, Schiller RM, Gischler SJ, van Heijst AF, Tibboel D, Aarsen FK, H IJ. Growing Up After Critical Illness: Verbal, Visual-Spatial, and Working Memory Problems in Neonatal Extracorporeal Membrane Oxygenation Survivors. Critical care medicine. 2016;44:1182–1190. doi: 10.1097/ccm.0000000000001626

7. Raets MM, Dudink J, Ijsselstijn H, van Heijst AF, Lequin MH, Houmes RJ, Wildschut ED, Reiss IK, Govaert P, Tibboel D. Brain injury associated with neonatal extracorporeal membrane oxygenation in the Netherlands: a nationwide evaluation spanning two decades. Pediatric critical care medicine : a journal of the Society of Critical Care Medicine and the World Federation of Pediatric Intensive and Critical Care Societies. 2013;14:884–892. doi: 10.1097/PCC.0b013e3182a555ac

8. Bembea MM, Felling RJ, Caprarola SD, Ng DK, Tekes A, Boyle K, Yiu A, Rizkalla N, Schwartz J, Everett AD, et al. Neurologic Outcomes in a Two-Center Cohort of Neonatal and Pediatric Patients Supported on Extracorporeal Membrane Oxygenation. ASAIO journal (American Society for Artificial Internal Organs : 1SS2). 2020;66:79–88. doi: 10.1097/mat.0000000000000933

9. Farhat A, Li X, Huet B, Tweed J, Morriss MC, Raman L. Routine Neuroimaging: Understanding Brain Injury in Pediatric Extracorporeal Membrane Oxygenation. Critical care medicine. 2022;50:480–490. doi: 10.1097/ccm.0000000000005308

10. Haydar B, Baetzel A, Elliott A, MacEachern M, Kamal A, Christensen R. Adverse Events During Intrahospital Transport of Critically Ill Children: A Systematic Review. Anesthesia and analgesia. 2020;131:1135–1145. doi: 10.1213/ane.0000000000004585

11. Knight PH, Maheshwari N, Hussain J, Scholl M, Hughes M, Papadimos TJ, Guo WA, Cipolla J, Stawicki SP, Latchana N. Complications during intrahospital transport of critically ill patients: Focus on risk identification and prevention. International journal of critical illness and injury science. 2015;5:256–264. doi: 10.4103/2229-5151.170840

12. Murata M, Nakagawa N, Kawasaki T, Yasuo S, Yoshida T, Ando K, Okamori S, Okada Y. Adverse events during intrahospital transport of critically ill patients: A systematic review and meta-analysis. The American journal of emergency medicine. 2022;52:13–19. doi: 10.1016/j.ajem.2021.11.021

13. Christy A, Murchison C, Wilson JL. Quick Brain Magnetic Resonance Imaging With Diffusion-Weighted Imaging as a First Imaging Modality in Pediatric Stroke. Pediatric neurology. 2018;78:55–60. doi: 10.1016/j.pediatrneurol.2017.09.020

14. McGlennan C, Ganesan V. Delays in investigation and management of acute arterial ischaemic stroke in children. Developmental medicine and child neurology. 2008;50:537–540. doi: 10.1111/j.1469-8749.2008.03012.x

15. Ko TS, Catennacio E, Shin SS, Stern J, Massey SL, Kilbaugh TJ, Hwang M. Advanced Neuromonitoring Modalities on the Horizon: Detection and Management of Acute Brain Injury in Children. Neurocritical care. 2023;38:791–811. doi: 10.1007/s12028-023-01690-9

16. Boyle K, Felling R, Yiu A, Battarjee W, Schwartz JM, Salorio C, Bembea MM. Neurologic Outcomes After Extracorporeal Membrane Oxygenation: A Systematic Review. Pediatric critical care medicine : a journal of the Society of Critical Care Medicine and the World Federation of Pediatric Intensive and Critical Care Societies. 2018;19:760–766. doi: 10.1097/pcc.0000000000001612

17. LaRovere KL, Vonberg FW, Prabhu SP, Kapur K, Harini C, Garcia-Jacques R, Chao JH, Akhondi-Asl A, Thiagarajan R, Tasker RC. Patterns of Head Computed Tomography Abnormalities During Pediatric Extracorporeal Membrane Oxygenation and Association With Outcomes. Pediatric neurology. 2017;73:64–70. doi: 10.1016/j.pediatrneurol.2017.05.006

18. Anton-Martin P, Braga B, Megison S, Journeycake J, Moreland J. Craniectomy and Traumatic Brain Injury in Children on Extracorporeal Membrane Oxygenation Support. Pediatric emergency care. 2018;34:e204–e210. doi: 10.1097/pec.0000000000000907

19. Ezetendu C, Baloglu O, Othman HF, Nandakumar V, Latifi S, Aly H. Stroke in pediatric ECMO patients: analysis of the National Inpatient Sample (NIS) database. Pediatric research. 2022;92:754–761. doi: 10.1038/s41390-022-02088-7

20. Golej J, Trittenwein G. Early detection of neurologic injury and issues of rehabilitation after pediatric cardiac extracorporeal membrane oxygenation. Artificial organs. 1999;23:1020–1025. doi: 10.1046/j.1525-1594.1999.06446.x

21. Beekman R, Crawford A, Mazurek MH, Prabhat AM, Chavva IR, Parasuram N, Kim N, Kim JA, Petersen N, de Havenon A, et al. Bedside monitoring of hypoxic ischemic brain injury using low-field, portable brain magnetic resonance imaging after cardiac arrest. Resuscitation. 2022;176:150–158. doi: 10.1016/j.resuscitation.2022.05.002

22. Mazurek MH, Cahn BA, Yuen MM, Prabhat AM, Chavva IR, Shah JT, Crawford AL, Welch EB, Rothberg J, Sacolick L, et al. Portable, bedside, low-field magnetic resonance imaging for evaluation of intracerebral hemorrhage. Nature communications. 2021;12:5119. doi: 10.1038/s41467-021-25441-6

23. Sheth KN, Mazurek MH, Yuen MM, Cahn BA, Shah JT, Ward A, Kim JA, Gilmore EJ, Falcone GJ, Petersen N, et al. Assessment of Brain Injury Using Portable, Low-Field Magnetic Resonance Imaging at the Bedside of Critically Ill Patients. JAMA neurology. 2020;78:41–47. doi: 10.1001/jamaneurol.2020.3263

24. Sheth KN, Yuen MM, Mazurek MH, Cahn BA, Prabhat AM, Salehi S, Shah JT, By S, Welch EB, Sofka M, et al. Bedside detection of intracranial midline shift using portable magnetic resonance imaging. Scientific reports. 2022;12:67. doi: 10.1038/s41598-021-03892-7

25. Yuen MM, Prabhat AM, Mazurek MH, Chavva IR, Crawford A, Cahn BA, Beekman R, Kim JA, Gobeske KT, Petersen NH, et al. Portable, low-field magnetic resonance imaging enables highly accessible and dynamic bedside evaluation of ischemic stroke. Science advances. 2022;8:eabm3952. doi: 10.1126/sciadv.abm3952

26. Cawley P, Padormo F, Cromb D, Almalbis J, Marenzana M, Teixeira R, Uus A, O’Muircheartaigh J, Williams SCR, Counsell SJ, et al. Development of neonatal-specific sequences for portable ultralow field magnetic resonance brain imaging: a prospective, single-centre, cohort study. EClinicalMedicine. 2023;65:102253. doi: 10.1016/j.eclinm.2023.102253

27. Sien ME, Robinson AL, Hu HH, Nitkin CR, Hall AS, Files MG, Artz NS, Pitts JT, Chan SS. Feasibility of and experience using a portable MRI scanner in the neonatal intensive care unit. Archives of disease in childhood Fetal and neonatal edition. 2023;108:45–50. doi: 10.1136/archdischild-2022-324200

28. Cho SM, Khanduja S, Wilcox C, Dinh K, Kim J, Kang JK, Chinedozi ID, Darby Z, Acton M, Rando H, et al. Clinical Use of Bedside Portable Low-field Brain Magnetic Resonance Imaging in Patients on ECMO: The Results from Multicenter SAFE MRI ECMO Study. Res Sq. 2024. doi: 10.21203/rs.3.rs-3858221/v1

29. Cho SM, Wilcox C, Keller S, Acton M, Rando H, Etchill E, Giuliano K, Bush EL, Sair HI, Pitts J, et al. Assessing the SAfety and FEasibility of bedside portable low-field brain Magnetic Resonance Imaging in patients on ECMO (SAFE-MRI ECMO study): study protocol and first case series experience. Critical care. 2022;26:119. doi: 10.1186/s13054-022-03990-6

30. Wilcox C, Acton M, Rando H, Keller S, Sair HI, Chinedozi I, Pitts J, Kim BS, Whitman G, Cho SM. Safety of Bedside Portable Low-Field Brain MRI in ECMO Patients Supported on Intra-Aortic Balloon Pump. Diagnostics (Basel). 2022;12. doi: 10.3390/diagnostics12112871

31. Sabir H, Kipfmueller F, Bagci S, Dresbach T, Grass T, Nitsch-Felsecker P, Pantazis C, Schmitt J, Schroeder L, Mueller A. Feasibility of bedside portable MRI in neonates and children during ECLS. Critical care. 2023;27:134. doi: 10.1186/s13054-023-04416-7

32. Cho S-M, Khanduja S, Wilcox C, Dinh K, Kim J, Kang J, Chinedozi I, Darby Z, Acton M, Rando H, et al. Clinical Use of Bedside Portable Ultra-Low-Field Brain Magnetic Resonance Imaging in Patients on Extracorporeal Membrane Oxygenation - Results From the Multicenter SAFE MRI ECMO Study. Circulation. 2024.

33. Bembea MM, Rizkalla N, Freedy J, Barasch N, Vaidya D, Pronovost PJ, Everett AD, Mueller G. Plasma Biomarkers of Brain Injury as Diagnostic Tools and Outcome Predictors After Extracorporeal Membrane Oxygenation. Critical care medicine. 2015;43:2202–2211. doi: 10.1097/ccm.0000000000001145

34. Lorusso R, Taccone FS, Belliato M, Delnoij T, Zanatta P, Cvetkovic M, Davidson M, Belohlavek J, Matta N, Davis C, et al. Brain monitoring in adult and pediatric ECMO patients: the importance of early and late assessments. Minerva anestesiologica. 2017;83:1061–1074. doi: 10.23736/s0375-9393.17.11911-5

35. O’Brien NF, Buttram SDW, Maa T, Lovett ME, Reuter-Rice K, LaRovere KL. Cerebrovascular Physiology During Pediatric Extracorporeal Membrane Oxygenation: A Multicenter Study Using Transcranial Doppler Ultrasonography. Pediatric critical care medicine : a journal of the Society of Critical Care Medicine and the World Federation of Pediatric Intensive and Critical Care Societies. 2019;20:178–186. doi: 10.1097/pcc.0000000000001778

36. Rilinger JF, Smith CM, deRegnier RAO, Goldstein JL, Mills MG, Reynolds M, Backer CL, Burrowes DM, Mehta P, Piantino J, et al. Transcranial Doppler Identification of Neurologic Injury during Pediatric Extracorporeal Membrane Oxygenation Therapy. Journal of stroke and cerebrovascular diseases : the official journal of National Stroke Association. 2017;26:2336–2345. doi: 10.1016/j.jstrokecerebrovasdis.2017.05.022

